# Meningoencephalitis associated with COVID-19: A systematic review

**DOI:** 10.1101/2020.06.25.20140145

**Authors:** Ritwick Mondal, Upasana Ganguly, Shramana Deb, Gourav Shome, Subhasish Pramanik, Deebya Bandyopadhyay, Durjoy Lahiri

## Abstract

**Background and aims:** With the growing number of COVID-19 cases in recent times, the varied range of presentations is progressively becoming an addressing issue among clinicians. A significant set of patients with extra pulmonary symptoms has been reported worldwide. Neurological involvement in the form of altered mental status, loss of consciousness in considerable amounts has drawn attention of physicians all across the globe. Here we venture out to summarise the clinical profile, investigations and radiological findings among patients with SARS-CoV-2 associated meningoencephalitis in the form of a systematic review, which may aid clinicians in early diagnosis and prognostic evaluation of the disease.

**Methodology:** This review was carried out based on the existing PRISMA (Preferred Report for Systemic Review and Meta analyses) consensus statement. The data for this review was collected from four databases: Pubmed/Medline, NIH Litcovid, Embase and Cochrane library and Preprint servers up till 10th June, 2020. Search strategy comprised of a range of keywords from relevant medical subject headings which includes “SARS-COV-2”, “COVID-19”, “meningoencephalitis” etc. All peer reviewed, case control, case report, pre print articles satisfying our inclusion criteria were involved in the study. The inclusion prerequisites comprised of confirmed SARS-CoV-2 cases with neurological manifestations, previous cases of SARS-CoV, MERS-CoV with neurological involvement provided all the studies were published in English language. Quantitative data was expressed in mean+/-SD, while the qualitative date in percentages. Paired t test was used for analysing the data based on differences between mean and respective values with a p value of <0.05 considered to be statistically significant.

**Results:** A total of 43 cases were involved from 24 studies after screening from databases and preprint servers, out of which 29 of them had completed investigation profile and were included in the final analysis. Clincial and Laboratory findings as well as neuroimaging findings (CT, MRI and MRS) revealed consistent presentations towards association of COVID-19 with meningoencephalitis. Epileptogenic pictures were also evident on EEG (electroencephalogram) findings.

**Conclusion:** SARS-CoV-2 has been isolated from CSF as well as cerebrum of cases with meningoencephalitis depicting the natural tendency of the virus to invade the central nervous system. Speculations about retrograde olfactory transport or alternative haematogenous spread seem to be correlating with above findings. This review may raise the index of suspicion about COVID-19 among clinicians while attending patients with neurological manifestations.

## 1- Introduction

The world at present is in combat with COVID-19, appropriately termed by Bill Gates as a ‘Once-in-a-Century’ Pandemic [**1**]. The dual challenge posed by this pandemic, namely saving lives as well as preventing horizontal transmission, has stretched clinicians to the extreme. With the passage of time, the situation has become more complicated given the unusual clinical manifestation of this viral infection. Several extra-pulmonary manifestations of COVID-19 have been brought to attention by treating physicians worldwide. Among these, neuro-invasive potential of SARS-CoV-2 has received significant attention. There is now sufficient body of evidence to support the idea that COVID-19 can have pure neurological presentations and on some occasions, preceding the typical respiratory manifestations [**2-4**]. Besides, a myriad of neurological consequences following typical clinical presentation of COVID-19 has also been documented across the globe. In sum, neuro COVID as a distinct topic of discussion has been steadily gaining attention over the last couple of months.

Among the central nervous system (CNS) manifestations, impaired consciousness/encephalopathy is a widely reported symptom of COVID-19 [**2**,**5**] and various possible etiologies underlying this symptom have been scrutinized in literature. It has been widely speculated that the virus gains access into the brain via olfactory bulb and the hypothesis gained substantial support from the observation that anosmia is a fairly consistent symptom of early COVID-19 [**2**]. An alternative proposed route for the virus to invade brain is hematogenous [**2**]. Given the potential of SARS-CoV-2 to invade CNS and also our previous experience with MERS and SARS-CoV, meningo-encephalitis in COVID-19 is a duly anticipated clinical feature. Indeed in the last couple of months, multiple reports of meningo-encephalitis associated with COVID-19 have surfaced up. These cases not only have encompassed a wide range of clinical presentations but also have documented varied laboratory and imaging results. That said, it can be assumed that in current situation under-reporting of cases is a non-negligible issue and therefore these cases only represent tip of the iceberg. Nevertheless, an organized summary and critical review of these documents can reveal a wealth of information about the clinical, laboratory and imaging features of meningo-encephalitis in COVID-19.

In this background, we set out to develop a systematic review of meningo-encephalitis cases in COVID-19 available in a wide array of databases. Our main objective is to summarize the clinical presentations, laboratory parameters including CSF abnormalities and brain imaging features of SARS-CoV-2 associated meningo-encephalitis. Such a documentation will not only work as a guide to clinicians dealing with COVID-19 patients, but also will open up new avenues towards understanding the neuro-invasive potential and routes of this virus.

## 2- Methodology

### 2.1- Design

This systematic review was conducted by following the Preferred Reporting for Systematic Review and Meta-Analysis (PRISMA) consensus statement (CRD42020185571) [**6**]. Studies relevant to the confirmed cases of COVID-19 infection with confirmed or suspected association of meningoencephalitis were included.

### 2.2- Search strategy

In this systematic review four databases:Pubmed/Medline, NIH LitCovid, Embase, and Cochrane Library were searched using pre specified searching strategies and this search was concluded on June 10, 2020. The search strategy consists of variation of keywords of relevant medical subject headings (MeSH) and key words, including “SARS-CoV-2”, “COVID-19”, “coronavirus”, “clinical symptoms”, “neurological impairments” and “Meningoencephalitis”. Severe Acute Respiratory Syndrome Coronavirus (SARS-CoV) and Middle East Respiratory Syndrome Coronavirus (MERS-CoV) were also included in our search strategy to capture related articles. We also hand searched additional COVID-19 specific articles using the reference list of the selected studies, relevant journal websites, and renowned pre-print servers (medRxiv, bioRxiv, pre-preints.org) from 2019 to current date for literature inclusion. To decrease publication bias, we invigilated the references of all studies potentially missed in electrical search. Content experts also searched the grey literature of any relevant articles.

### 2.3- Study selection criteria

All peer-reviewed, pre-print (not-peer-reviewed) including cohort, case-control studies and case reports which met the pre-specified inclusion and exclusion criteria were included in this study.

### 2.4- Inclusion criteria

Studies met the following inclusion criteria were included if: (i) Conducted for the patients infected with COVID-19 with or suspected meningoencephalitis (ii) Studies registering neurological manifestations of COVID-19 patients were included with encephalitis like symptoms (iii) Parallel studies to look into the detailed distribution and incidences of meningoencephalitis in previous outbreaks i.e. SARS-CoV, MERS-CoV and various other coronaviruses were compared with current pandemic in the discussion section(iv) published in the English language. Studies without complete information but met our inclusion criteria were included in the narrative review.

### 2.5- Exclusion Criteria

Studies excluded if COVID-19 was not confirmed among patients and written in languages other than English. We also excluded review papers, viewpoints, commentaries, and studies where no information related to neurological symptoms or meningoencephalitis was reported.

### 2.6- Data extraction

Prior to the screening process, team of three reviewers (GS, DB and SD) participated in calibration and screening exercises. First two reviewers (GS and DB) subsequently screened independently the titles and abstracts of all identified citations, and the third reviewer (SD) verified those citations and screened papers by (GS and DB). Other two reviewers (UG and SA) then retrieved and screened independently the full texts of all citations deemed eligible by the reviewer (SD) on the team and analysed those data. Another reviewer (RM) independently verified these extracted full texts for eligibility towards analysis and designed the overall study structure. The corresponding author (DL) had resolved disagreements whenever necessary and took final decisions regarding the study. Throughout the screening and data extraction process, the reviewers used piloted forms. In addition to the relevant clinical data, the reviewers also extracted data on the following characteristics: Study characteristics (i.e. study identifier, study design, setting, timeframe); population characteristics; comparator characteristics, outcomes (qualitative and/or quantitative); Clinical factors (definition and measurement methods); reported funding sources and conflict of interests; study limitations. The Newcastle-Ottawa scale was used to assess the selection procedure, the comparability and the outcomes of each reviewed study.

### 2.7- Statistical analysis

Quantitative data were presented using means ± standard deviation (SDs).Qualitative data were presented as percentage value. Unit discordance among the variables was resolved by converting the variables to a standard unit of measurement. Further, one sample t-test was performed to find out significant difference between mean value and respective reference value (highest value of the reference range was taken) of each parameter. A value of ‘p’ < 0.05 was considered as statistically significant. All statistical analyses were performed and analyzed using Graph pad prism (Version 6, Sandiego, CA, USA). A meta analysis was planned to analyze the association of the demographic findings, co-morbidities, symptoms, diagnostic parameters and outcomes with imaging findings but was later omitted due to lack of sufficient data.

## 3- Results

### 3.1- Study characteristics-(Table-1)

A total of around 43 cases were included in the study from 24 different articles coming from both database search and preprint servers. Complete data were available for 29[n=29] cases and laboratory findings data was unavailable for rest of 14 cases from the paper of Andrea Pilloto et al. The studies of 29 cases comprised of case reports and original article.

**Figure 1.**
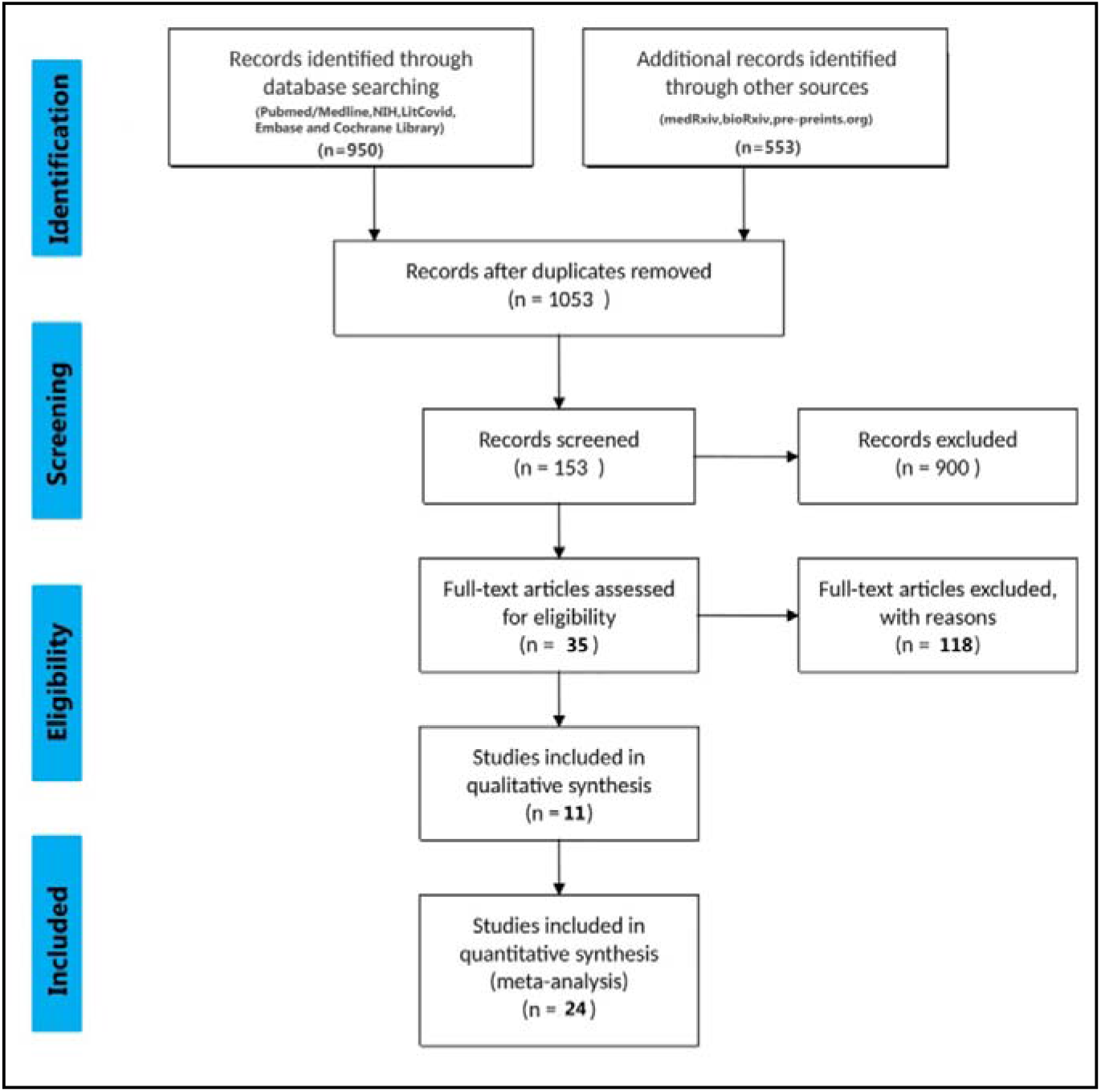

**TABLE 1.** Included Studies

| SL.No | Authors | Title of the paper | No. of cases | Type of Paper |
| --- | --- | --- | --- | --- |
| 1. | Po Fung Wong et al<br>(UK) | A case of rhombencephalitis as a rare complication of acute COVID-19 infection | 1 | Lessons of the Month |
| 2. | Adelaide Panariello et al<br>(Italy) | Anti-NMDA receptor encephalitis in a psychiatric Covid-19 patient: A case report | 1 | Letter to Editor |
| 3. | Mohammad Al Olama et al<br>(UAE) | COVID-19-associated meningoencephalitis complicated with intracranial hemorrhage: a case report | 1 | Case Report |
| 4. | H. Chaumont et al<br>(France) | Acute meningoencephalitis in a patient with COVID-19 | 1 | Letter to Editor |
| 5. | Andrea Pilotto et al<br>(Italy) | COVID-19 impact on consecutive neurological patients admitted to the emergency department | 14 | Original article |
| 6. | Rong Yin et al<br>(China) | Concomitant neurological symptoms observed in a patient diagnosed with coronavirus disease 2019 | 1 | Letter to Editor |
| 7. | Gary N. McAbee et al<br>(USA) | Encephalitis Associated with COVID-19 Infection in an 11 Year-Old Child | 1 | Letter to Editor |
| 8. | Sandeep Sohal, Mansoor Mosammat<br>(USA) | COVID-19 Presenting with Seizures | 1 | Original Article |
| 9. | Rebecca Packwood et al<br>(USA) | An Unusual Case Report of COVID-19 Presenting with Meningitis Symptoms and Shingles | 1 | Case Report |
| 10. | Charcon Aguilar et al<br>(Spain) | COVID-19: Fever syndrome and neurological symptoms in a neonate | 1 | Letter to Editor |
| 11. | Hale Afshar et al<br>(Iran) | Evolution and resolution of brain involvement associated with SARS-CoV2 infection: A close Clinical – Paraclinical follow up study of a case | 1 | Case Report |
| 12. | Ye et al<br>(China) | Encephalitis as a clinical manifestation of COVID-19 | 1 | Letter to Editor |
| 13. | Narges Karimi et al<br>(Iran) | Frequent Convulsive Seizures in an Adult Patient with COVID-19: A Case Report | 1 | Case Report |
| 14. | Ibrahim Efekan Efe<br>(Turkey, Germany) | COVID-19-associated encephalitis mimicking glial tumor: a case report | 1 | Case Report |
| 15. | Duong et al<br>(USA) | Meningoencephalitis without respiratory failure in a young female patient with COVID-19 infection in Downtown Los Angeles, early April 2020 | 1 | Letter to Editor |
| 16. | Moriguchi et al<br>(Japan) | A first case of meningitis/encephalitis associated with SARS-Coronavirus-2 | 1 | Case Report |
| 17. | Misayo Hayashi et al<br>(Japan) | COVID-19-associated mild encephalitis/encephalopathy with a reversible splenial lesion | 1 | Letter to Editor |
| 18. | Andrea Pilotto et al<br>(Italy) | Steroid-responsive encephalitis in Covid-19 disease | 1 | Original Article |
| 19. | Dogan et al<br>(Turkey) | Plasmapheresis treatment in COVID-19-related autoimmune meningoencephalitis: Case series | 6 | Original Article |
| 20. | Debaleena Mukherjee et al<br>(India) | Ataxia as a presenting manifestation of COVID -19: Report of a single case | 1 | Case Report |
| 21. | Raphael Bernard-Valnet et al<br>(Switzerland) | Two patients with acute meningo-encephalitis concomitant to SARS-CoV-2 infection | 2 | Letters to Editor |
| 22. | Mauro Morassi et al<br>(Italy) | Stroke in patients with SARS-CoV-2 infection: case series | 1 | Original Article |
| 23. | Guy Talmor et al<br>(USA) | Nasoseptal Flap Necrosis After Endoscopic Skull Base Surgery in the Setting of SARS-CoV-2/COVID-19 | 1 | Case Report |
| 24. | Manuel Romero et al<br>(Spain) | Neurologic manifestations in hospitalized patients in COVID-19 The ALBACOVID registry | 1 | Original Article |

### 3.2- Demography and Clinical Symptomatology-(Table-2A and Table 2B)

The selected study comprised of 29 cases with the mean age around (50.8±19.09) and 20 (70%) male with 9(30%) female cases were reported for analysis with full data availability. SARS-CoV-2 was primarily detected in nasopharyngeal swab (86%), CSF (10%) and antibody detected in two patients (7%) only. Majority of the reported cases had no travel history (7%) or contact with COVID-19 positive individuals (14%). They had various symptoms ranging from fever, cough, headache etc. Potent neurological symptoms reported were tonic-clonic seizures (10%), disorientation to time & place (14%), nuchal rigidity (17%), limb ataxia (21%), etc. Various kinds of co-morbidities that existed in patients were diabetes mellitus (21%), hypertension(28%), Obesity(7%). coronary artery disease (7%) with two special cases of Alzheimer’s disease and autism .The patients were hospitalized on (8.88±4.094) days (Mean ± SD) from the onset of the symptoms. Their average days of hospitalization were (13.83±7.901) days (Mean ± SD).

**TABLE 2A.** Demographic characteristics of patients hospitalized with COVID-19

| **Demographic feature** | **Total**<br>**(n=29)** |
| --- | --- |
| **Age (Years, Mean ± SD)** | 50.8±19.09 |
| **Sex (n, [%])**<br>*Male*<br>*Female* | 20 (70%)<br>9 (30%) |
| **Social history (n, [%])**<br>*Travel history*<br>*Contact with Covid-positive individuals* | 2 (7%)<br>4 (14%) |
| **Systemic Co-morbidities (n, [%])**<br>*Diabetes mellitus*<br>*Hypertension*<br>*Obesity*<br>*Coronary Artery Disease*<br>*Chronic Kidney Disease*<br>*Cancer*<br>*Substance Abuse*<br>*Cerebrovascular Disease*<br>*Hyperlipidemia*<br>*Autism*<br>*Alzheimer's Disease* | 6(21%)<br>8(28%)<br>2(7%)<br>2(7%)<br>1(3.5%)<br>1(3.5%)<br>1(3.5%)<br>1(3.5%)<br>1(3.5%)<br>1(3.5%)<br>1(3.5%) |
| **SARS – Cov2 detection (n, [%])**<br>*CSF*<br>*Nasopharyngeal swab*<br>*Antibody* | 3(10%)<br>25(86%)<br>2(7%) |
| **Days between symptom appearance and hospital admission (Mean ± SD)**<br>**Days of hospitalization (Days, Mean ± SD)** | 8.88±4.094<br>13.83±7.901 |

**Note**
n=no. of cases for which data was available for that particular variable
Total number of cases = 29

**TABLE 2B.** Clinical signs and symptoms

| **Symptoms** | **Total**<br>**(n=29)** |
| --- | --- |
| **General Symptoms (n, [%])** |  |
| *Fever* | 17(59%) |
| *Cough* | 8(28%) |
| *Fatigue* | 3(10%) |
| *Headache* | 7(24%) |
| *Breathing difficulty/Shortness of*<br>*breath* | 4(14%) |
| *Anosmia* | 1(3.5%) |
| *Ageusia* | 1(3.5%) |
| *Dysgeusia* | 1(3.5%) |
| *Myalgia* | 5(17%) |
| *Nausea* | 1(3.5%) |
| *Dizziness* | 3(10%) |
| *Diarrhea* | 4(14%) |
| *Anorexia* | 1(3.5%) |
| *Rhinorrhea* | 2(7%) |
| *Vomitting* | 2(7%) |
| *Abdominal pain* | 1(3.5%) |
| *Constipation* | 1(3.5%) |
| **Neurological symptoms (n, [%])** |  |
| *Tonic-clonic seizures* | 3(10%) |
| *Confusion/Disorientation /Altered*<br>*HMF* | 12(41.37%) |
| *Verbal and motor perseverations* | 1(3.5%) |
| *Bilateral grasping* | 2(7%) |
| *Visual hallucinations* | 2(7%) |
| *Nuchal rigidity* | 5(17%) |
| *Speech slurring* | 3(10%) |
| *Kernig's sign* | 1(3.5%) |
| *Babinski sign* | 1(3.5%) |
| *Chaddock sign* | 1(3.5%) |
| *Brudzinski sign* | 2(7%) |
| *Limb ataxia* | 6(21%) |
| *Increased deep tendon reflexes* | 1(3.5%) |

**Note**
n=no. of cases for which data was available for that particular variable
Total number of cases = 29

**TABLE 2C.** Clinical and Laboratory diagnostic parameters

| Test | Normal value | Mean ± SD | p value |
| --- | --- | --- | --- |
| **Blood pressure (mm Hg) [n=5]** |  |  |  |
| *Systolic Pressure* | 120 | 128.2±22.68 | 0.4641 |
| *Diastolic Pressure* | 80 | 78.60±9.12 | 0.3513 |
| **Heart rate (beats/min) [n=5]** | 82 | 93±4.41 | **0.004\*** |
| **O<sub>2</sub> saturation [at room air] (%) [n=8]** | 97 | 93.63±3.88 | **0.0120\*** |
| **Respiratory rate (breaths per minute) [n=8]** | 20 | 25.33±7.448 | 13.98 |
| **WBC [n=13]** | (4-11)$\times 10^9$ | (10.12±8.33)$\times 10^9$ | 0.734 |
| **Lymphocyte [n=6]** | (1.5-3.5) $\times 10^9$ | (3.177±3.06) $\times 10^9$ | 0.8717 |
| **Platelets [n=6]** | (150-450) $\times 10^9$ | (326.4±258.8) $\times 10^9$ | 0.345 |
| **C-reactive protein (mg/L) [n=13]** | <10 mg/L | (102 ± 129.8) | 0.064 |
| **D-dimer (ng/ml) [n=10]** | 500 ng/ml | (3970±3217) | **0.0186\*** |
| **LDH (U/L) [n=13]** | 140-280 | 642±494.9 | 0.059 |
| **CSF parameters** |  |  |  |
| *Protein (mg/dl)[n=18]* | 15-45 | 73.61±56.31 | **0.00457\*** |
| *Glucose (mg/dl)[n=16]* | 45-80 | 95.24±42.16 | 0.2170 |
| *Lymphocyte [n=6]* | 62% | 95.33±5.68 | **0.009\*** |
| *IgG (mg/L)[n=5]* | 0-81 | 4.91±1.32 | **<0.0001\*** |
| *IgG index (mg/L)[n=5]* | 0-0.7 | 1.50±1.84 | 0.4479 |
| *AlbQ [n=5]* | 1.85 | 10.32±3.65 | **<0.0001\*** |

**Note-**
n=no. of cases for which data was available for that particular variable
Total number of cases = 29
\* indicates p<0.05 which is considered statistically significant

### 3.3- Laboratory Findings

#### 3.3.1 Hematological parameters-(Table-2C)

Major cell counts were observed within various patients. Recorded WBC counts in cases[n=13] were (10.12±8.33)×10^9^ Mean ± SD) with a p value of (p=0.734) and Lymphocyte counts observed for six patients[n=6] were (3.177±3.06) ×10^9^ Mean ± SD). Elevated levels of C-reactive protein among patients [n=13] around (102 ± 129.8 Mean ± SD)(mg/L) were reported with a p value of 0.064. Moreover highly significant levels of D-dimer (ng/ml) in patients [n=10] were observed with a significant p value of 0.0186. (p<0.05)

#### 3.3.2 CSF parameters – (Table-2C)

Various CSF parameters under biochemical analysis such as Glucose(mg/dl) [n=16], Protein(mg/dl) [n=18] were recorded where significant levels of protein (p=0.00457)with elevation of (73.61±56.31) mg/dl from the normal range of (15-45)mg/dl was observed .Moreover certain cell count parameters such as Lymphocyte [n=6] was significantly (p=0.009) higher than the normal range.. Antibody like I_g_G (mg/L) was also found in significant (p=0.0001) amount in five patients [n=5].

#### 3.3.3 EEG findings – (Table-3)

EEG findings were reported in 9 cases and in the remaining 20 cases this parameter was not available for analysis. Bernard Valnet et al reported EEG from one patient as abundant bursts of anterior low-medium voltage irregular spike- and waves superimposed on an irregularly slowed theta background [**27**]. Whereas Sohal *et al* documented that the patient had six left temporal seizures, left temporal sharp waves which were epileptogenic in nature [**14**]. Pilotto *et al* showed generalized slowing with decreased reactivity to acoustic stimuli [**24**] and bilateral slowed activity without seizures was reported by Chaumont *et al* [**10**]. Generalized slowing with no epileptic discharges was also documented by Duong et al [**21**]. McAbee *et al* only reported Frontal intermittent delta activity [**13**]. On the other hand Adelaide Panariello *et al* reported Theta activity at 6 Hz, unstable and non reactive to visual stimuli without significant asymmetries [**8**]. Continuous monitoring with amplitude-integrated electroencephalography (EEG) by Charcon Aguilar *et al* for 36 hours revealed a continuous background pattern with sleep-wake cycles in the absence of electrical and clinical seizures for a patient [**16**]. Interestingly Morassi *et al* reported that on day 4 the EEG showed a normal background in the alpha range (8 Hz) associated with recurrent sharp slow waves over the left temporal region, which occasionally were seen also on the right homologous regions. On day 10, a new EEG excluded non-convulsive status epilepticus, while showing persistence of sharp slow waves, mainly over the left hemispheric regions for the same patient [**28**].

**TABLE 3.**
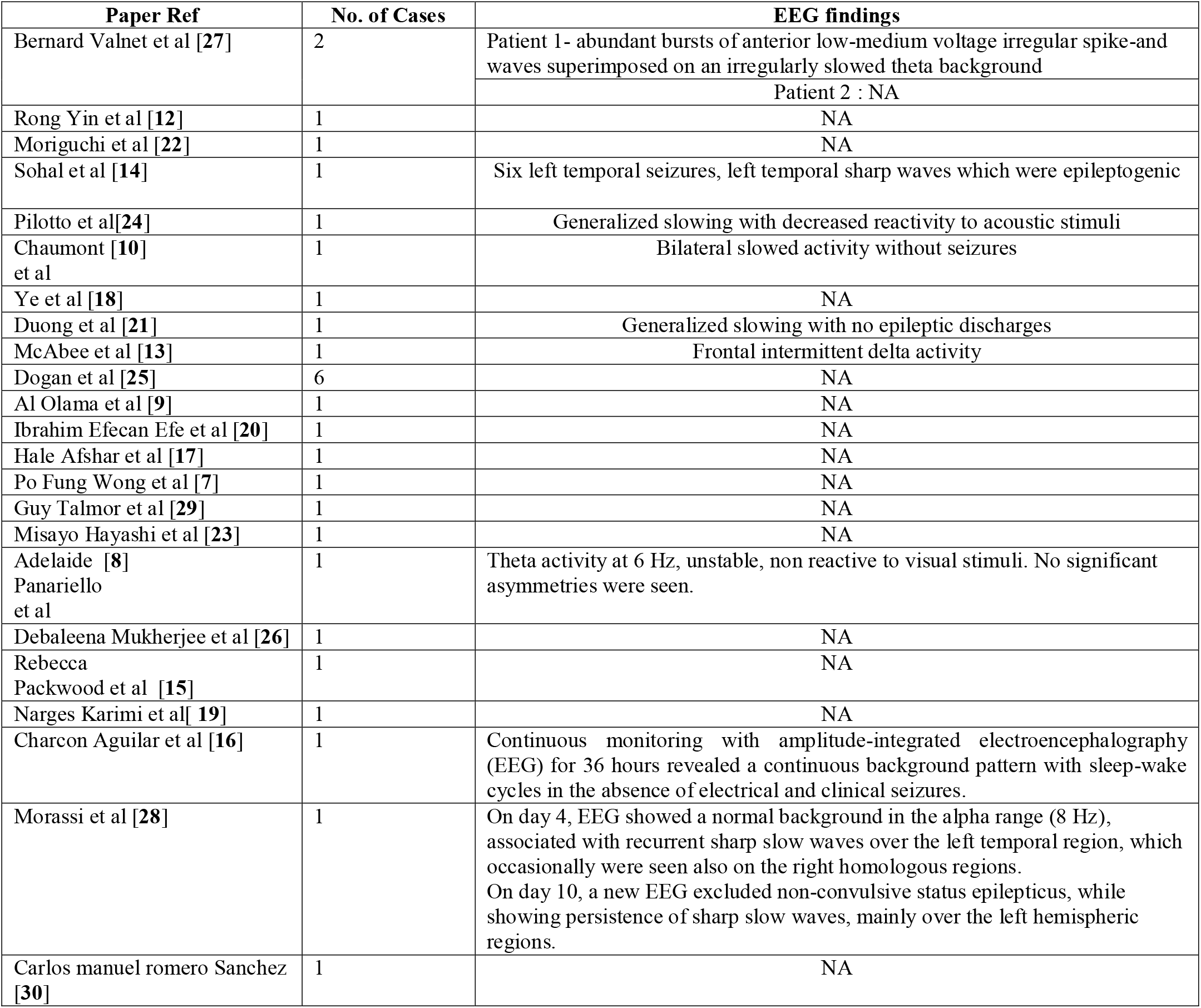
EEG Findings

#### 3.3.4 Neuroimaging findings-(Table-4)

Neuroimaging findings were reported in 20 studies consisting of 26 cases and in the remaining 3 cases brain scans were not available. Among the imaging modalities, CT scan and MRI were done in 10 and 16 cases respectively. Vessel imaging was available in 1 case (CTA). In majority of the cases neuroimaging finding did not reveal any abnormality (13/26, 50%). At least one radiological abnormality was reported in 14 cases (14/26, 53.84%). Among the abnormal MRI findings, T2/T2 FLAIR hyperintensity in MRI brain was the most frequently reported 6/16, 37.5%). Noteworthy, temporal lobe was the commonest site of involvement in MRI brain (6 cases). Stroke like findings manifested by hypodensity on CT scan (3 cases) and diffusion restriction in MRI (2 cases) were also observed. Beaded appearance on CT angiography was reported in 1 case associated with extradural hemorrhage.

**TABLE 4.** Neuroimaging findings

| SL.No | Imaging Type | Findings |
| --- | --- | --- |
| 1. | CT | **a. NAD-** [ref-8,12,13,18,21,24,26]<br>**b. Hemorrhagic-**<br>Frontal EDH [ref-9]<br>**c. Ischemic-**<br>Right caudate nucleus [ref-28]<br>Chronic microvascular change [ref-14] |
| 2. | MRI | **a. NAD-**[ref-10,19,25 (case no 3,4,5),27]<br>**b. T<sub>2</sub>** [ref-7,17,20] **or T<sub>2</sub> /FLAIR hyperintensity**-[ref-20,22,28,30]<br>**c. Encephalitis like feature**<br>**(hyperintensity,enhancement,hemorrhage)** [ref-25( case no1,2,6)]<br>**d. DWI Restriction/ADC hypodensity** [ref-22,23] |
| 3. | Angiography | **a. CTA**<br>Beaded appearance-[ref-9]<br>**b. MRA**<br>NA |
| 5. | MRS | **a. Choline peak**-[ref-20] |
| 4. | Not Available | [ref-11,15,16] |

**Abbreviations-**
- Computed Tomography (CT) - Magnetic Resonance Imaging (MRI) - Magnetic Resonance Spectroscopy (MRS) - No Appreciable Disease (NAD) - Fluid Attenuated Inversion Recovery (FLAIR) hyperintensity - Diffusion-Weighted Imaging- (DWI) - Apparent diffusion Coefficient (ADC) - Computed Tomography Angiography (CTA) - Magnetic Resonance Angiography (MRA)

## 4- Treatment and outcome-(Table-5)

Treatment were carried out with various drugs such as Hydroxycholoroquine (52%),Azithromycin (34%), Lopinavir/Ritonavir (14%), Favipiravir(28%) etc or used in multidrug therapy(66%). Various procedures such as Plasmapheresis(21%), invasive ventilation (34%), were used along with many other therapies on the patients .Frequently used multidrug therapy includes Hydroxycholorquine along with Azithromycin(55.17%). Majority of the patients recovered (61%) with a mortality of (17%) and six patients were still under treatment during the period of study.

**TABLE 5.** Treatment and Outcome

|  |  |
| --- | --- |
| **Combination Therapy** |  |
| *Multidrug Therapy* | 19(66%) |
| *Most common multidrug*<br>*(Hydroxychloroquine and*<br>*Azithromycin)* | 16(55.17%) |
| **Therapeutics** |  |
| *Hydroxychloroquine (HCQ)* | 15(52%) |
| *Azithromycin (AZT)* | 10(34%) |
| *Remdesivir* | 1(3.5%) |
| *Lopinavir/Ritonavir (LOP/RIT)* | 4(14%) |
| *Favipiravir (FAV)* | 8(28%) |
| *Ceftriaxone (CEF)* | 5(17%) |
| *Ganciclovir* | 1(3.5%) |
| *Meropenem* | 2(7%) |
| *Amoxicillin* | 4(14%) |
| *Acyclovir* | 7(24%) |
| *Arbidol* | 1(3.5%) |
| *Ribavirin* | 1(3.5%) |
| *Vancomycin* | 4(14%) |
| *Darunavir/Cobisitat* | 1(3.5%) |
| *Plasmapheresis* | 6(21%) |
| *Invasive ventilation* | 10(34%) |
| *IVIG* | 2(7%) |
| *Steroids* | 5(17%) |
| *Mannitol infusion* | 1(3.5%) |
| **Outcome** |  |
| *Recovery rate* | 18(62%) |
| *Still in treatment* | 6(21%) |
| *Mortality rate* | 5 (17%) |

## 5- Discussion

In the index paper we attempted to summarize the clinical features, laboratory parameters, EEG findings and imaging abnormalities in patients presenting with confirmed SARS-CoV-2 infection and clinical / confirmed meningo-encephalitis. Of note, a fair number of cases with meningo-encephalitic presentation of COVID-19 have already surfaced up even in the background of possible under-reporting because of the ongoing pandemic. We found description of 43 such cases in the available literature so far among which 29 were included in the final analysis as detailed data for the remaining 14 cases were not available. To the best of our knowledge, this is the first ever attempt to explore meningo-encephalitis in COVID-19 by means of systematic review of both peer reviewed as well as pre-print data. The present study is important in terms of advancing our knowledge and understanding related to the neuro-invasive potential of SARS-CoV-2 which primarly is considered a respiratory pathogen

Coronaviruses have been detected in both cerebrum and cerebrospinal fluid of individuals with seizures, encephalitis, and encephalomyelitis[**31**].According to different case reports of MERS coronavirus, neurological manifestations such as meningoencephalitis, hyporeflexia, ataxia have been found[**32**]. It was also shown that neuronal infection leading to death in hACE2(full form) transegnic mice infected with SARS-CoV [**33**]. Accumulative evidences reflect that virus has been detected in cerebrum[**34**] as well as in CSF in case of SARS-CoV infection[**35**]. Other coronaviruses such as HCoV-OC43 under the genera of *Betacoronaviridae*,is found to be associated with viral encephalitis [**36**], acute disseminated encephalomyelitis[**37**].Similarly,CoV-NL63 under the genera of *Alphacoronaviridae* causes acute encephalitis with self limitng CNS infection[**38**].Viral encephalitis caused by coronavirus infection was reported among pediatric population with respiratory tract infection [**39**].Overall the predisposition of various coronaviruses towards neurotropism seems evident and very much happening which is generating a major concern for virus mediated encephalitis.

Analysis of the demographic parameters reveals that mean age of the patients with COVID-19 related meningo-encephalitis was 50.8 (±19.09) years with males being more commonly affected (70%). Mean latency between symptom onset and hospital admission was close to 9 days while mean number of days of hospital stay was approximately 14 days. Majority of the documented patients had co-morbidities with diabetes mellitus and hypertension being most frequent. Among the general symptoms, fever was by far the commonest followed by cough. Notably, anosmia and ageusia (commonly considered as markers of neuro-invasion) were reported in uncommonly. In contrast, diarrhea was relatively common (14%) among the general symptoms. Respiratory distress was reported in 4 (14%) of the documented cases.

Various neurological symptoms were reported in mening-encephalitis associated with SARS-CoV-2 infection. Confusion or disorientation to time and place or altered mental status was the most frequently reported symptom accounting for 41.37% of the cases. While the classic Kernig’s and Brudzinski’s signs were not frequently documented, nuchal rigidity have been more commonly (17%) observed. Limb ataxia was reported in more than 1/5^th^ of the cases including the case reported by one of the authors of the present review. Other less reported neurological symptoms include tonic clonic seizures, slurred speech and cognitive difficulties such as verbal and motor perseveration. SARS-CoV-2 detection in CSF was reported in only 3 cases whereas majority (86%) of the patients were diagnosed based on nasopharyngeal swab testing.

Among the clinical signs pertaining to general survey, tachycardia and reduced oxygen saturation were common findings. Analysis of blood parameters reveal raised CRP, D-dimer and LDH were prominent findings while occurrence of lymphopenia or leucopenia did not reach statistical significance. CSF parameters often give valuable insight to the treating clinicians when faced with cases of meningo-encephalitis. CSF parameters in COVID-19 associated meningo-encephalitis raised protein and increased lymphocyte count as the most notable observations. Therefore it can be assumed that brain infection in the background of SARS-CoV-2 infection may lead to CSF pleocytosis with lymphocytic predominance accompanied by significant level of rise in CSF protein, both pointing towards intra-thecal inflammation. In addition, while there was documented dip in CSF IgG levels, the IgG index did not deviate much from the normal limits; which may be a reflection of hypoglobulinemia resulting from lymphopenia [40].

Among the abnormal EEG findings documented in COVID-19 encephalitis, focal epileptiform discharges and generalized slowing are most notable. While focal discharges may signify the occurrence of focal convulsions, generalized slowing is commensurate with the high frequency of altered mental status observed thus far.

Majority of the patient’s brain scans were found devoid of any abnormality. However, among those with abnormal brain scans, a wide range of structural lesions have been observed. The commonest abnormality reported was T2/FLAIR hyper-intensity affecting the temporal lobes. Among the viral causes of temporal lobar hyperintensity, HSV is the forerunner followed by CMV while anti-NMDAR related autoimmune encephalitis frequently presents temporal lobar changes [41]. In addition, stroke like presentations have also been noted which are characterized by restricted diffusion in DWI.

As far as the treatment is concerned, more than 50% patients have received therapy with HCQ while significant proportion of patients received azithromycin, favipiravir and acyclovir. In fact, almost all the patients were on multi-drug regimen as was documented in this review. Reported mortality rate in COVID-19 encephalitis is 17%. Majority (62%) of the affected patients experienced recovery and around 1/5^th^ were still under treatment at the time of drafting this review.

## 6- Conclusion

In this systematic review we observe, quite a few reports of meningo-encepalitis associated with COVID-19 are already available in recent literature even in the face of possible under-reporting in contrast to the earlier epidemics of MERS and SARS-CoV. Various clinical presentations have surfaced up among which confusion or disorientation is the most frequent. Among the laboratory parameters, raised CRP and D-dimer are most prominent findings. Analysis of CSF parameters reveals, high protein and lymphocytic pleocytosis are the most observable abnormalities. Focal epileptiform discharges and slowing of background are common EEG abnormalities observed in COVID-19 related meningo-encephalitis. A myriad of radiological abnormalities have been observed of which temporal lobar hyper-intensity and diffusion restriction are common. This review has attempted to provide a basic outline of COVID-19 related meningo-encephalitis from a clinician’s perspective. Hopefully with increment in reporting of neurological manifestations of COVID-19, a clearer picture will become available towards the future.

## Data Availability

The data for this systematic review is available freely

## 7- Acknowledgement

We are sincerely thankful to Rohan Sarkhel (Department of Computer Science Engineering,Maulana Abul Kalam Azad University,India) for preparing illustration for this paper.

